# Automated Design of Patient-Specific 4D-Printed Phantoms for Quality Assurance of Adaptive Radiotherapy on a 1.5T MR-Linac

**DOI:** 10.64898/2026.07.09.26357659

**Authors:** Horatio Montero Hamkins, Ka Ho Tam, Angela Sobremonte, Sandeep Jogi, Eugene Koay, Comron Hassanzadeh, Paul Segars, Neelam Tyagi, Ergys Subashi

## Abstract

**Background:** Independent end-to-end verification of adaptive radiotherapy on MR-Linac systems is limited by the lack of patient-specific phantoms able to reproduce imaging and dosimetric properties from CT and MRI scanners. We present a method for automated generation of 4D, patient-specific, multi-material 3D-printable phantoms for quality assurance of adaptive radiotherapy on a 1.5T MR-Linac.

**Methods:** Patient images were automatically segmented using a pretrained deep learning model. The segmented structures were converted into high-resolution 3D meshes and assembled into printable phantoms. A dosimeter holder was inserted at user-defined anatomical locations, with orientation optimized to avoid traversal across heterogeneous tissue interfaces. Physiological motion was incorporated by generating phantoms from images at different timepoints and interpolating deformation fields to create continuous 4D models. Multi-material organs designed by mixing a set of six polymers at various proportions were used to reproduce tissue-specific imaging properties. The properties of material mixtures were evaluated in a clinical CT simulator and a 1.5T MR-Linac.

**Results:** The proposed workflow enables automated generation of anatomically realistic phantoms with several types of embedded dosimeters. A discrete search method was designed for placement and immobilization of OSLD, film, and ion chamber dosimeters. Calibration curves for Hounsfield units were derived through variations in radiopaque material content, while MR signal intensity was modulated by gel and tissue matrix mixtures. Patient-derived abdominal phantoms were fabricated at multiple scales while replicating internal anatomical detail. Multi-dimensional phantom generation enabled continuous representation of motion states with consistent mesh topology across phases.

**Conclusions:** We demonstrate an end-to-end workflow for automated generation of 4D patient-specific phantoms for MR-Linac quality assurance. The method combines realistic anatomy, embedded dosimetry, multimodal imaging properties, and physiological motion within a single fabrication framework. This approachmay enable an improved validation of adaptive radiotherapy workflows in MR-guided treatment devices.

## INTRODUCTION

Online and offline adaptive radiotherapy enabled by high-field MR-Linac systems represents a major advance in precision radiation delivery, allowing real-time visualization of patient anatomy and daily plan adaptation [1–4]. However, this capability introduces new challenges for quality assurance (QA), particularly in the presence of magnetic-field-induced dose perturbations such as the electron return effect (ERE) and electron streaming effect (ESE) [5–7]. These effects introduce spatial dose heterogeneities at tissue interfaces and along magnetic field lines, necessitating validation measurements that extend beyond those found in conventional linac QA methods [8].

An independent end-to-end validation of adaptive workflows requires phantoms capable of reproducing patient-specific anatomy, imaging characteristics across both CT and MRI scanners, and representative dosimetric properties under magnetic field conditions. Existing QA tools and methods - including homogeneous phantoms, detector arrays, gel dosimeters, and Monte Carlo simulations - provide valuable but often incomplete representations of the clinical setup. Homogeneous phantoms lack realistic anatomical modeling, gel dosimeters are challenging to handle and calibrate for routine clinical use, and Monte Carlo (MC) simulations, while accurate, do not provide a physical validation platform for integrated imaging, planning, and delivery systems [9–11].

Recent work has explored both digital and physical phantoms for MR-Linac QA [12–14]. Virtual patient-specific phantoms and log-file-based dose reconstruction methods have been proposed for verification of adaptive workflows [10,15,16]. Similar to MC simulations, these approaches do not fully capture the coupled imaging-dosimetry-delivery chain in a physical system. Additionally, studies evaluating magnetic field effects using specialized QA devices or simplified geometries have highlighted the complexity of ERE/ESE but lack anatomical fidelity required for patient-specific validation.

Advances in 3D printing have enabled the development of anthropomorphic phantoms with increasing anatomical detail and material properties. Multiple fabrication techniques - including fused deposition modeling, stereolithography, polymer jetting, and selective laser sintering - have been investigated for radiological phantoms [17–22]. Additive manufacturing technologies offer distinct advantages due to high spatial resolution 3D printing and ability to mix multiple materials at the voxel level. In the context of QA for MR-Linacs, this enables independent tuning of mechanical and dosimetric properties, CT Hounsfield units, and MR signal characteristics within a single printed object. Recent studies have demonstrated the feasibility of multi-material phantoms for CT and MRI applications, including the use of radiopaque additives to modulate attenuation and soft polymer mixtures to replicate MR contrast [23–25]. However, most existing implementations remain limited to static geometries and do not incorporate patient-specific anatomy, embedded dosimetry, or physiological motion.

Digital phantoms such as the extended cardiac-torso (XCAT) phantom present a framework for simulating anatomical motion, including respiratory, cardiac, and gastrointestinal motility [26,27]. While these models enable detailed 4D simulations, translation to patient-specific physical phantoms suitable for end-to-end QA remains limited.

In this work, we present a method for generating 4D patient-specific, multi-material 3D-printable phantoms for QA of adaptive radiotherapy on a 1.5T MR-Linac. The proposed framework integrates deep learning–based segmentation, high-resolution mesh generation, automated dosimeter positioning, and multi-material printing with calibrated CT and MR properties. Furthermore, physiological motion is incorporated through temporal interpolation of anatomically consistent meshes, enabling continuous 4D phantom representations. This approach aims to provide a practical and scalable solution for end-to-end validation of adaptive radiotherapy workflows in MR-guided devices. This work represents a design and methodological framework, with our initial validation focused specifically on material characteristics, and demonstration of workflow feasibility rather than active clinical deployment.

## MATERIALS AND METHODS

### Overview and Digital Phantom Development

An automated workflow was developed to generate patient-specific, multi-material 3D-printable phantoms from clinical imaging data for quality assurance of adaptive radiotherapy on a 1.5T MR-Linac. The workflow was designed to support both static and time-resolved image datasets and to incorporate user-defined dosimetry targets, automated anatomical modeling, multi-material fabrication, and physiological motion.

An initial proof-of-concept development was performed using the XCAT phantom, which provided a digital reference for establishing the core steps of the pipeline, including anatomical grouping, mesh generation, dosimeter path design, and preparation for fabrication. The anatomical structures defined in the XCAT reference model were grouped into the following organ systems relevant for radiotherapy applications: lung, esophagus, heart, blood vessels, stomach, liver, gallbladder, pancreas, small bowel, large bowel, kidneys, bones, spinal cord, skin, connective tissue, and muscles. Structures not explicitly included in this set were accounted for using a global body mask representing the full patient volume. For radiotherapy of static targets away from the abdominothoracic cavity, the default organ grouping in XCAT was not changed.

For each anatomical group, binary volumetric masks were generated and converted into stereolithography (STL) surface meshes using a MATLAB (Mathworks, Natick, MA) implementation of the marching cubes algorithm. The resulting STL objects were imported into Meshmixer v3.5 (Autodesk, San Francisco, CA) for additional pre-processing and mesh editing. The goal of pre-processing and editing was to minimize layering artifacts (resulting from stacked image slices) and open surfaces (for partially imaged structures). These were corrected by solidification and remeshing operations to produce watertight geometries suitable for Boolean subtraction and 3D printing. The ‘Make Solid’ tool was used with the following parameters: Solid type = accurate, Solid accuracy = 512, Mesh Density = 512. Surface smoothing was subsequently performed (smoothing type = shape preserving, smoothing scale = 20) to reduce voxelization artifacts while preserving anatomical shape. Representative renderings of the resulting XCAT anatomical systems are shown in Fig. 1.

**Figure 1.**
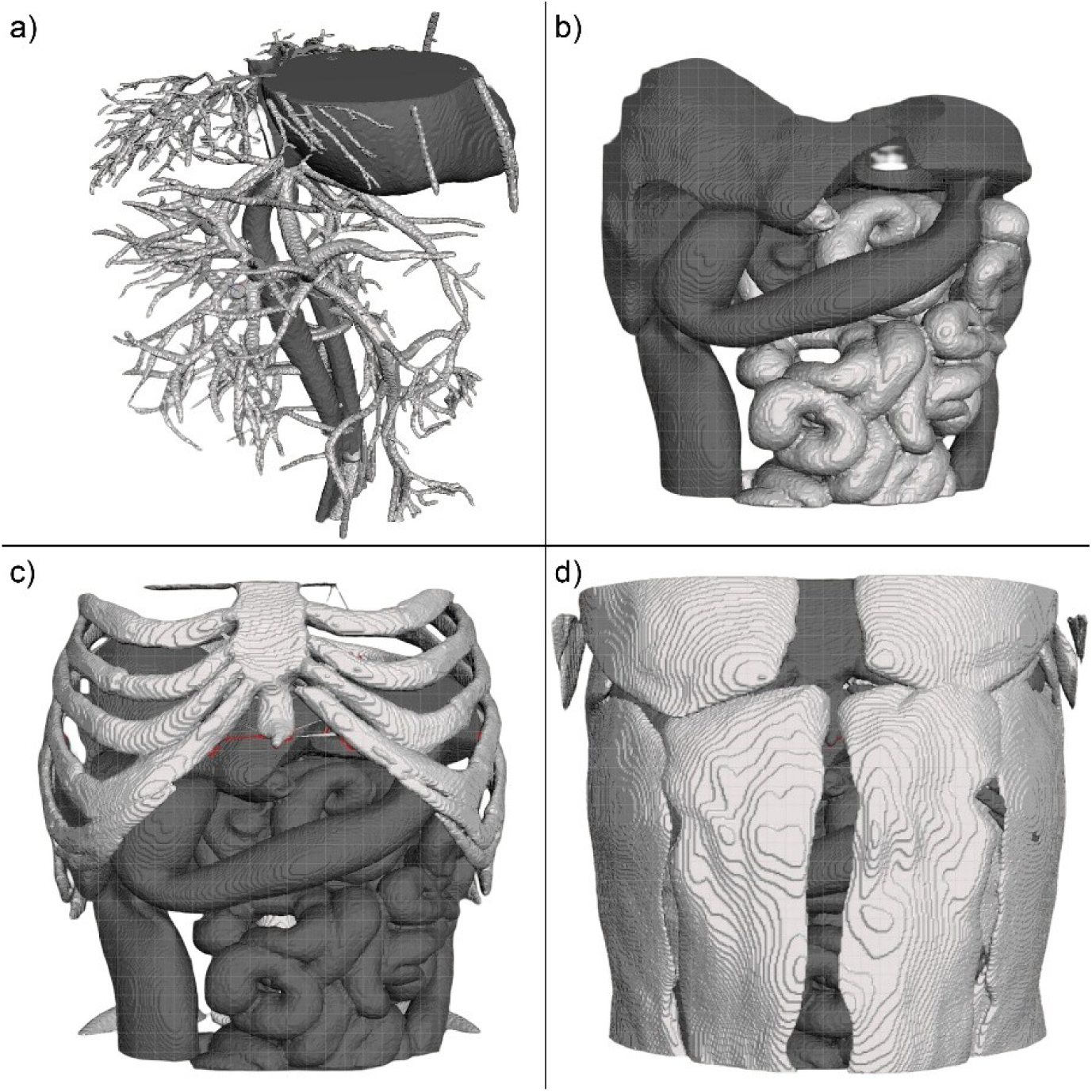
Surface rendering of anatomical systems displaying (a) cardiovascular system, (b) gastrointestinal system, (c) skeletal system overlaid on the gastrointestinal system, (d) muscular system overlaid on the combined anatomy.

This initial workflow established the geometric framework later extended to patient-specific CT and MR datasets. In the final pipeline, the process begins in the treatment planning system (TPS), where the user selects a voxel or region of interest (ROI) for dosimetric measurements. The selected coordinates, together with the patient imaging data, are then exported for automated phantom generation.

### Patient-specific phantom generation

To extend the workflow from digital phantom proof-of-concept to patient-specific phantom generation, pretrained deep learning-based segmentation was integrated into the pipeline. Patient CT or MR images were automatically segmented into anatomical structures, then converted into printable 3D models. The segmentation pipeline used TotalSegmentator v2.10.0 [28,29] installed and executed locally to maintain compatibility with clinical data privacy requirements and to enable automated batch processing. The segmentation output consisted of binary volumetric masks stored in NIfTI format, which were converted into 3D NumPy arrays and subsequently transformed into surface meshes using a marching cubes implementation in Python v3.13.

Compared with the initial XCAT-based workflow, the patient-specific implementation replaced the segmentation-derived anatomical groups with direct structure extraction from the patient image. Each segmented organ was represented as an independent 3D mesh, allowing downstream material assignment and phantom assembly on an organ-by-organ basis. A body mask was again used to represent the remaining patient volume and to provide a continuous external boundary for fabrication.

Geometry processing, assembly, and dosimeter placement were automated using Python scripts and the Blender Python API. Blender v4.5 was used in place of manual mesh-editing software so that Boolean operations, object tracking, and dosimeter placement could be performed programmatically. To support reproducible automation, a template with a structured file hierarchy and naming convention was implemented to track the original images, segmented structures, mesh files, dosimeter objects, and intermediate Boolean outputs throughout the workflow.

### Dosimeter placement, holder design, and phantom assembly

A central objective of the workflow was to enable accurate point or planar dosimetry within anatomically realistic phantoms. To achieve this, a dosimeter holder was automatically inserted into a given phantom at a user-defined target location. A Boolean subtraction was performed to create a physical cavity extending from the exterior surface of the phantom to the selected measurement point. This process is illustrated in Fig. 2.

**Figure 2.**
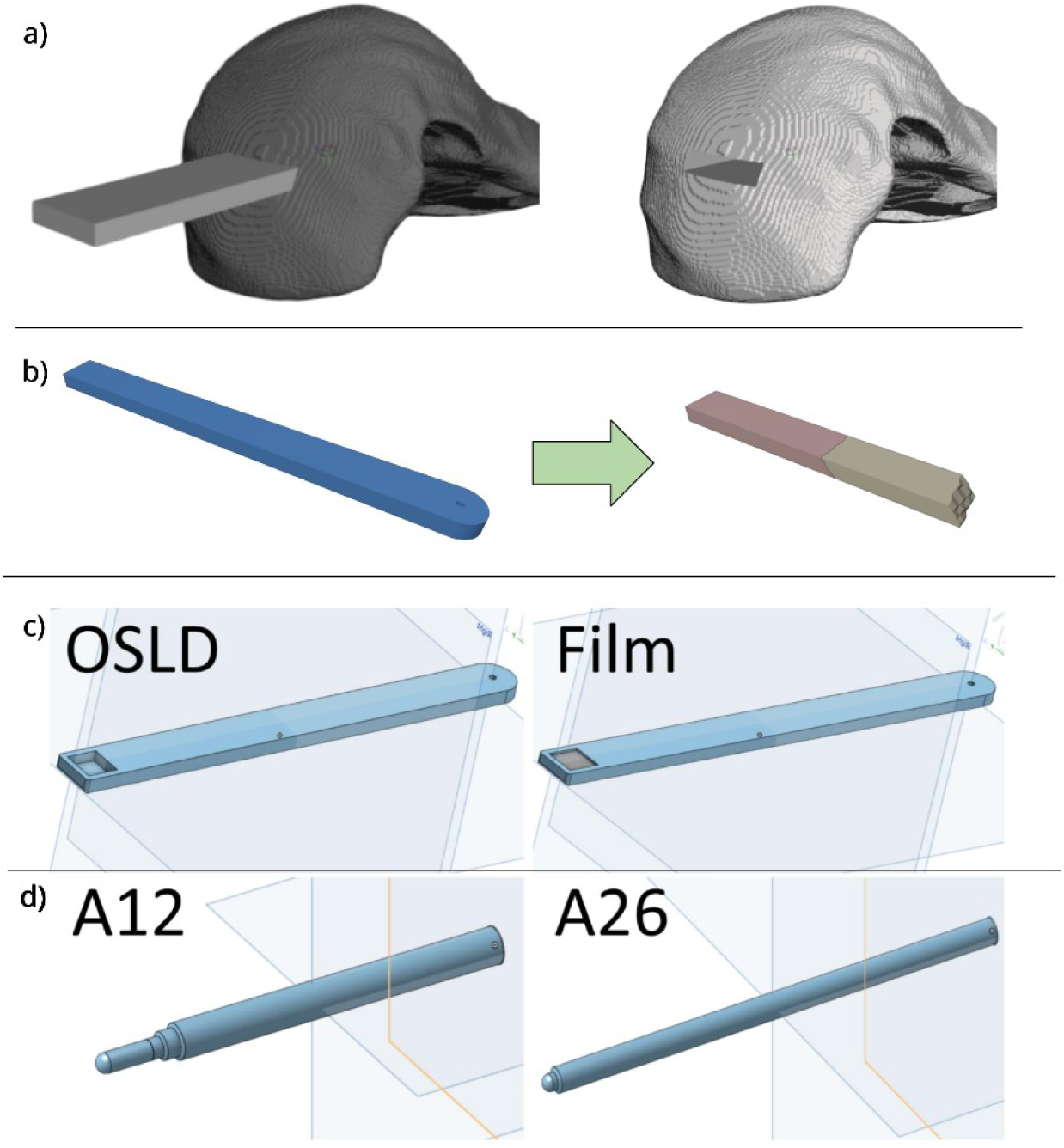
Schematic diagram of the process for dosimeter placement, material selection for the dosimeter holder, and available dosimeter models. (a) Dosimeter and holder inserted in the organ of interest and Boolean operator to create path for dosimeter holder (b) Initial template of dosimeter holder and demonstration of its material composition to match anatomy it transverses, showing two materials only (c) Holders for OSLD and film dosimetry (d) Holders for models of ionization chambers of various sensitive volumes. OSLD=Optically Stimulated Luminescence Dosimeter

The insertion path was designed to minimize perturbation of the local phantom composition and to preserve geometric fidelity after fabrication. In particular, the dosimeter orientation was selected to minimize intersections with anatomical structures having large differences in Hounsfield units or MR signal intensity. Reducing such intersections decreases the number of material transitions required in the holder and limits discontinuities along the insertion path. In addition, shorter insertion paths were preferred when possible, because longer holders are susceptible to bending or buckling during insertion, which may increase contact forces against the surrounding phantom, induce local shearing, and geometric misalignment.

In the trial phase using the XCAT-based workflow, this path was determined manually. In the automated patient-specific implementation, dosimeter placement and orientation were optimized programmatically. The ROI selected in the TPS was exported as Cartesian coordinates, and a discrete search algorithm was used to evaluate candidate holder orientations. For this proof-of-concept implementation, the holder was (i) centered on the ROI, (ii) placed along the anterior-posterior direction, (iii) rotated in 45° increments along the axial plane thus yielding eight candidate orientations. For each orientation, the algorithm evaluated the intersections between the holder and surrounding anatomical structures and selected an insertion trajectory that minimized the number and extent of such intersections.

Custom holders were designed for several dosimeter types, including optically stimulated luminescent dosimeters (OSLDs), radiochromic film, and ionization chambers. Holders for OSLDs and film were designed as fully enclosed top-and-bottom assemblies. Ion chamber holders were designed as hollow and partial enclosures with an extended outer support, allowing the chamber to be inserted directly and secured in place. The designs are shown in Fig. 2(c-d).

To reduce disruption of the local phantom properties, we partitioned the holder into material regions corresponding to the tissues it traversed. For example, the portion of the holder inside a target organ was assigned material properties matching that organ, whereas the extra-cavitary portion was assigned properties consistent with the surrounding body mask. This multi-material partitioning improved integration of the holder into the phantom and reduced abrupt material discontinuities at the holder-phantom boundaries.

After dosimeter placement, all organ meshes were subtracted from the body mask to remove overlaps and eliminate empty gaps between structures. Final remeshing and smoothing operations (octree_depth = 5, smoothing scale = 20) were then applied to produce a non-intersecting composite geometry suitable for fabrication.

### Multi-material fabrication and 4D motion modeling

Each assembled phantom is prepared for fabrication using a multi-material PolyJet 3D (Stratasys, Minnetonka, MN) printing system. The system allows for voxel-level mixing of printable materials, enabling spatial tuning of physical and imaging properties across anatomical structures. To approximate patient-specific tissue properties, mixtures of GelMatrix™, TissueMatrix™, BoneMatrix™, Agilus30™, Vero PureWhite™, and RadioMatrix™ were evaluated. We hypothesized that RadioMatrix™ can be used primarily to modulate radiographic attenuation, whereas gel and tissue matrix materials can be used to tune MR signal characteristics.

To characterize the imaging properties of candidate material mixtures, a series of calibration samples was fabricated in 5cm x 5cm x 5cm cubes and imaged using a clinical CT simulator (Siemens Healthineers, Erlangen, Germany) and a 1.5T MR-Linac (Elekta Inc., Stockholm, Sweden). Material combinations were systematically varied to establish the relationship between composition and CT or MR signal intensity. The tested combinations are summarized in Appendix A and Appendix B. All images were reconstructed using a voxel size of 1mm x 1mm x 1mm. CT images were acquired at a tube current of 100 mAs and four tube voltages: 80 kVp, 100 kVp, 120 kVp, 140 kVp. A 3D turbo-spin echo MRI sequence was used for T2w imaging with TR/TE=1300/87 ms. A 3D fast-field echo MRI sequence was used for T1w imaging with TR/TE=11/4.6 ms.

To incorporate physiological motion, the workflow was extended from static anatomical models to phantom models at various motion states. For temporally-resolved image data, a phantom was generated independently for each motion state. A reference body mesh corresponding to largest volume in the 4D volume was used to maintain consistent topology across the time series. The reference was used to constrict the volume of mesh pre-processing and editing across time steps. Before generating motion, non-manifold edges in each mesh were closed and a remeshing operation was performed (octree_depth = 5) limiting the object’s resolution to a 32×32×32 grid of voxels. Continuous motion was then generated by interpolating vertex positions between successive phases, producing a 4D phantom representation with anatomically smooth and consistent motion states. This process enabled interpolation in both spatial and temporal domains and allowed phantoms to be generated at arbitrary time points beyond the temporal resolution of the original image acquisition. Additionally, this framework supports generation of phantoms representing respiratory, cardiac, gastrointestinal, or combined physiological motion, while maintaining compatibility with patient-specific anatomy, embedded dosimetry, and multi-material fabrication. A static interpolated phantom may then be used for QA of gated radiotherapy. The 4D phantom with physiological motion may be used for 4D dosage calculation. Respiratory phase resolved images were acquired using clinical sequences in the CT/MRI scanners and sorted into 10 phases based on amplitude of respiratory signal.

## RESULTS

### Dosimeter holder design and phantom assembly

The dosimeter-placement framework enabled generation of insertion paths from the phantom surface to user-defined anatomical targets. As shown in Fig. 2(a-b), Boolean subtraction was used to create a cavity conforming to the holder geometry, while the holder itself was partitioned into regions corresponding to the tissues traversed along the insertion path. This design allowed the holder to blend with surrounding phantom structures while maintaining access to the selected dosimetry location.

The same framework supported multiple detector configurations. Representative holder designs are shown for OSLD and film dosimetry in Fig. 2(c), and for ion chamber models A12 and A26 (Standard Imaging, Middleton, WI) in Fig. 2(d). These results demonstrate that the insertion and holder-generation workflow can accommodate both planar and point dosimetry within the same design pipeline.

### Automated patient-specific phantom generation

A representative patient-specific model derived from a T2-weighted MRI is shown in Fig. 3. The user-selected dosimetry target is indicated within the liver on the source image in Fig. 3(a). After automated structure extraction and mesh generation, Boolean subtraction was used to create a dosimeter cavity at the selected location, as shown in Fig. 3(b). The resulting composite model preserved the relevant anatomy surrounding the target and provided a printable assembly for patient-specific phantom construction, as shown in Fig. 3(c).

**Figure 3.**
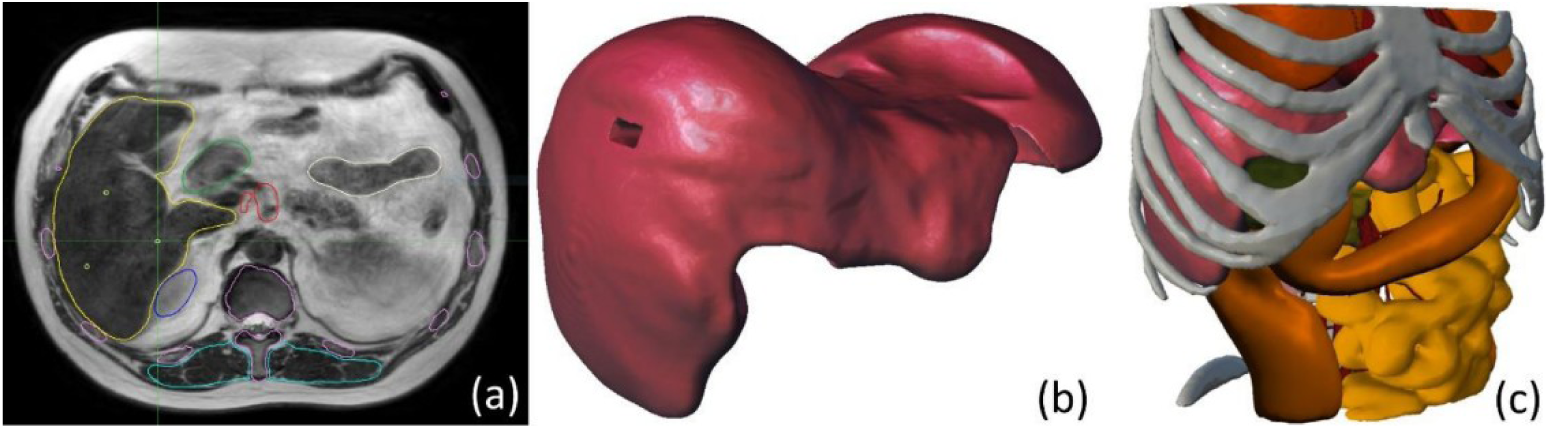
Representative model derived from a T2-weighted MRI of patient anatomy. (a) Axial slice from T2-weighted MRI displaying at the cross-hair the location of the dosimeter placement, (b) Model of the liver after the Boolean operation for dosimeter placement at the selected location, (c) Combined model representing the anatomy of interest.

A schematic overview of the automated patient-specific workflow is shown in Fig. 4. After image export from the treatment planning system, patient structures were segmented automatically, converted into 3D object files (.obj), and assembled into a composite phantom. The user-selected dosimetry target was then used as the endpoint for automated dosimeter placement. Together, Fig. 3 and Fig. 4 demonstrate the progression from clinical imaging and target selection to a dosimeter-ready, patient-specific phantom geometry.

**Figure 4.**
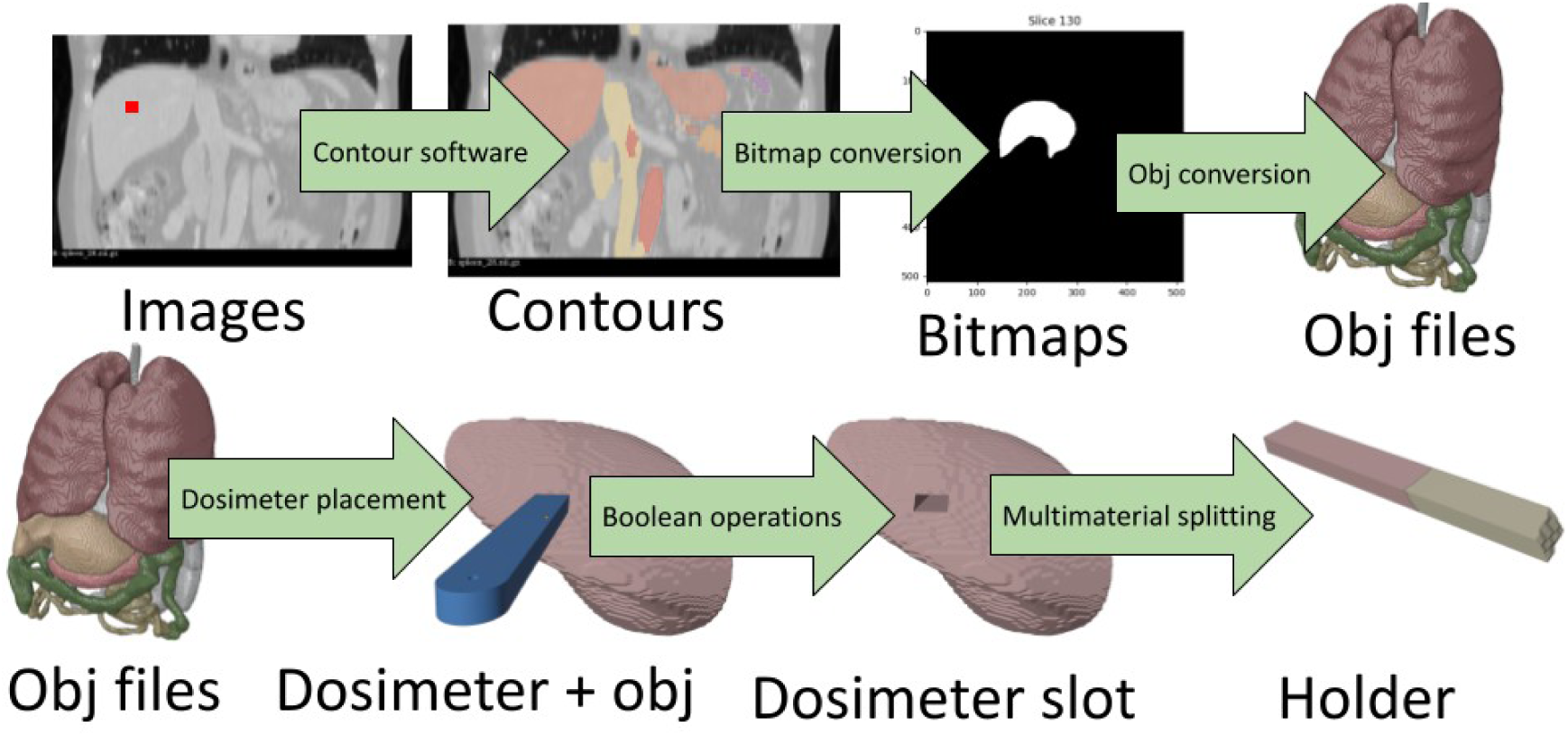
Step-by-step process for automated patient specific phantom design. Users select in the treatment planning system a region-of-interest for dose measurements. Patient images and location of dose measurements are then exported, contoured automatically, and converted into 3D .obj files. Finally, a user-specified dosimeter and its holder are placed at the target location and material specifications are determined for the holder.

### 4D phantom generation

The workflow was extended from static phantom design to generation of temporally resolved 4D phantoms. As illustrated in the XCAT phantom in Fig. 5, time-resolved images were separated into distinct motion states, and an independent phantom was generated for each state using the same automated pipeline applied to the static case. This allowed the anatomical structures present at each phase to be segmented, converted into 3D meshes, and incorporated into a phase-specific phantom representation. For patient specific phantom generation, the workflow is applied to 4D patient images.

**Figure 5.**
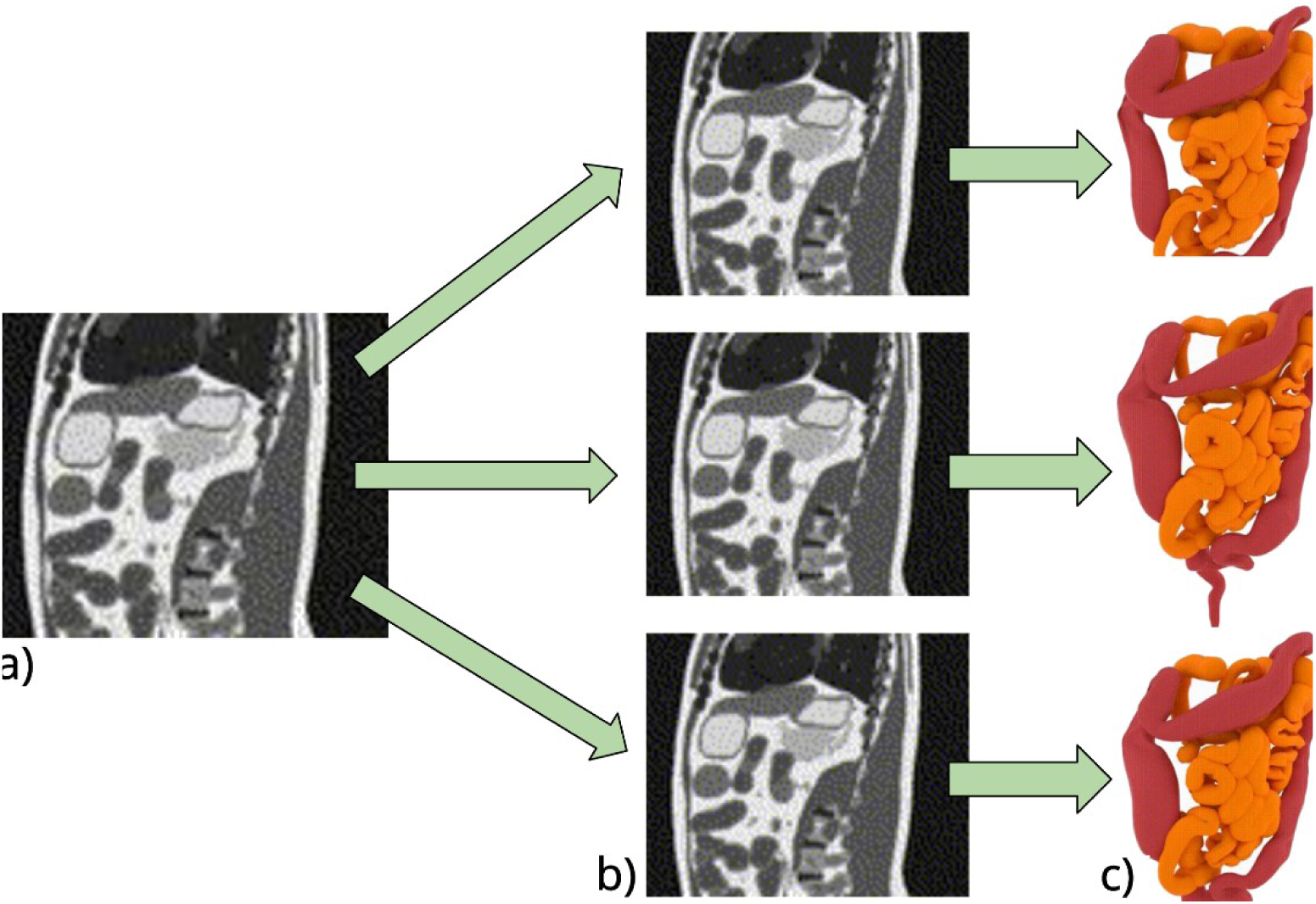
Visualization of 4D phantom generation process in the XCAT phantom. (a) Time-resolved images are split into (b) different motion states. Phantoms are then generated automatically for each motion state included in the 4D patient image.

To maintain anatomical consistency across time, a reference mesh was propagated through the motion states so that corresponding structures preserved a common topology from phase to phase. This was important for enabling smooth temporal transitions and avoiding discontinuities that would arise if each phase was treated as an independent surface model. Thereafter, vertex positions were interpolated between successive phases to generate intermediate motion states not explicitly present in the original image acquisition. As a result, the method produced continuous 4D phantom representations with smooth anatomical deformations. A rendering of a patient-specific phantom is included as a movie in Supplementary Materials.

This framework allowed interpolation in both the spatial and temporal domain, enabling phantom generation at arbitrary points beyond the spatiotemporal sampling of the original dataset. In practical terms, the method provides a way to model dynamic anatomy with higher effective temporal resolution than that of the source images while preserving the patient-specific geometry defined by the acquired data. The resulting 4D phantom therefore captures anatomical structure and the evolution of that structure through physiological motion. Also, given that the same geometric framework was used in each motion state, the 4D implementation remained compatible with downstream dosimeter placement and multi-material fabrication. The approach is therefore applicable to respiratory motion, gastrointestinal motion, cardiac motion, or combinations, and provides a basis for designing motion-aware phantoms for adaptive radiotherapy QA on MR-Linac systems.

### Material characterization and representative printed phantoms

Material characterization demonstrated that the imaging properties of the printed phantoms could be systematically tuned through controlled mixing, which allowed for the generation of modality-specific calibration curves.

For CT, it was found that the percentage of RadioMatrix™ can be varied to generate a range of Hounsfield units. The measurements in Fig. 6(a) show that the HU values depended on both material composition and tube voltage, providing a practical calibration framework for selecting mixtures that approximate the attenuation properties of patient organs. In particular, increasing radiopaque content expanded the achievable HU range, allowing materials to be selected according to the anatomical structure being modeled.

For MRI, it was found that combinations of GelMatrix™ and TissueMatrix™ were most effective at modulating signal intensity in T1- and T2-weighted imaging. The corresponding measurements in Fig. 6(b) show that material composition influenced signal intensity differently across contrast mechanisms, supporting the feasibility of tuning phantom materials for MR-visible anatomy. Rather than providing only geometric rigidity, the printed structures could also be designed to generate distinct MR contrast, which is particularly relevant for phantoms intended for MR-guided adaptive radiotherapy workflows.

The calibration curves in Fig. 6(a-b) collectively show that CT and MR properties can be adjusted through different aspects of the material formulation. Radiographic attenuation and MR signal characteristics were not controlled by a single mixture parameter, but by complementary material components. This separation is useful for multi-material phantom construction, because it supports a flexible assignment of imaging properties across organs and surrounding tissues.

Representative fabricated phantoms modeling abdominal anatomy at different scales are shown in Fig. 6(c-d). These examples demonstrate that the patient-derived meshes could be translated into physically realizable multi-material prints while preserving overall anatomical form and internal structural detail and rigidity. The printed models also show that the workflow is not restricted to a single phantom size, but can be adapted to different scales depending on the intended QA application.

**Figure 6.**
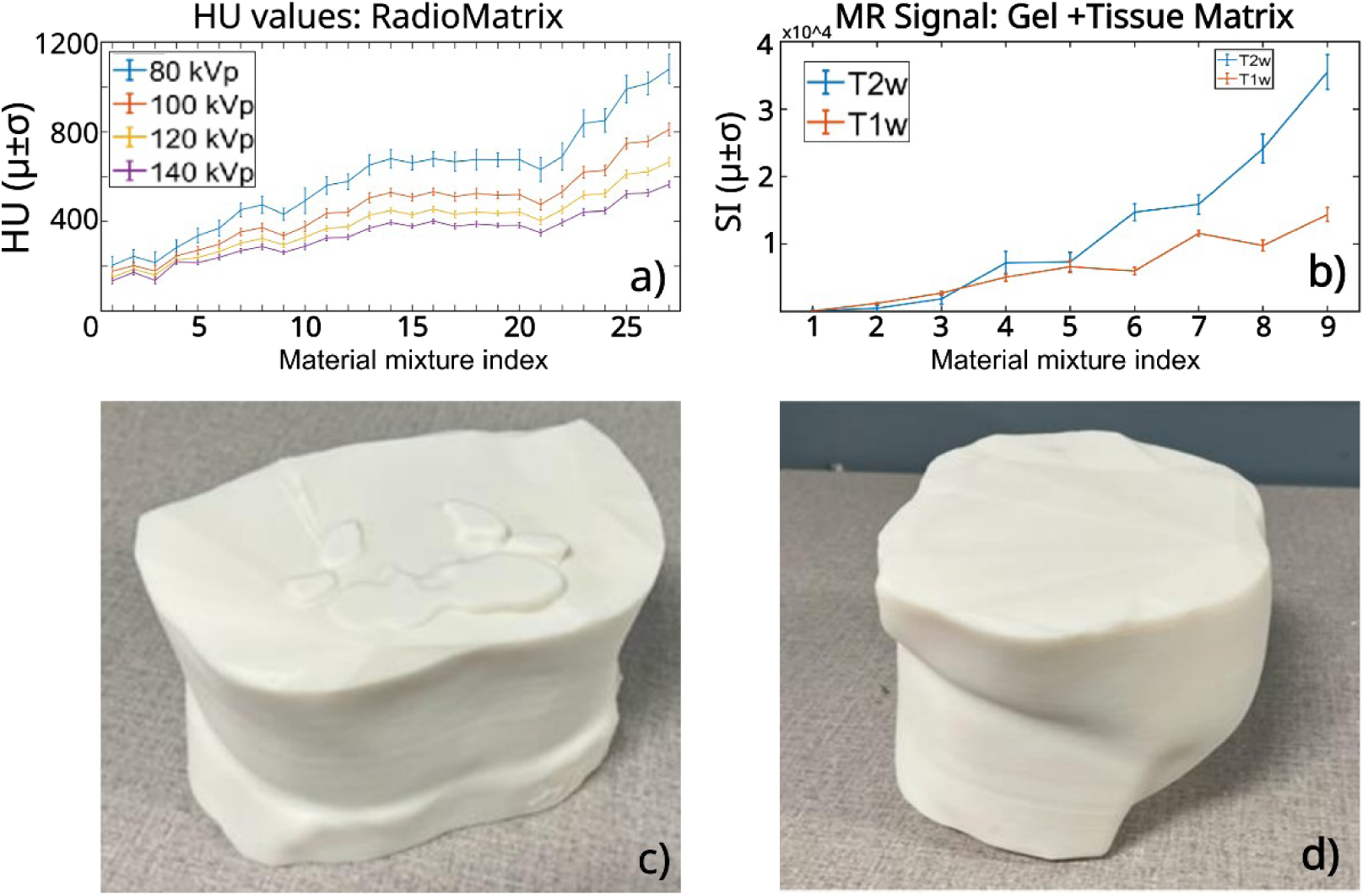
Calibration curves for material composition in phantoms designed for computed tomography or magnetic resonance imaging. (a) RadioMatrix™ present in material sample is increased from 10-100% shows that Hounsfield values are a function of material and tube voltage. (b) Signal intensity as a function of material and MRI contrast mechanism (T1 vs T2), as % composition of GelMatrix is increased from 10-60%. (c,d) Representative patient-specific phantoms modelling abdominal anatomy at various scales. HU=Hounsfield units, SI=signal intensity, T1w/T2w=T1/T2-weighted

## DISCUSSION

This work presents an end-to-end framework for automated design of patient-specific 4D-printed phantoms for quality assurance of adaptive radiotherapy on a 1.5T MR-Linac. A design and methodological framework was introduced with our initial validation focused specifically on material characteristics, and demonstration of workflow feasibility rather than active clinical deployment. The proposed method integrates automated image segmentation, mesh generation, dosimeter-holder placement, multi-material assignment, and motion modeling. In this proof-of-principle study, we find that this approach can generate realistic phantom geometries from patient images, support multiple dosimeter configurations, and produce multi-material phantoms with tunable CT and MR imaging properties.

The main contribution of this work is the combination of several capabilities that are typically addressed separately. Existing QA tools can support selected aspects of MR-Linac verification, but usually do not provide patient-specific anatomy, multimodal imaging contrast, embedded dosimetry, and physiological motion within a common device. In contrast, the workflow presented here begins with clinical imaging data and produces a phantom design that is compatible with both fabrication and downstream dosimetric measurements. This is particularly relevant for adaptive radiotherapy, where QA must account not only for dose calculation and delivery, but also for the coupled effects of anatomy, imaging, and workflow decisions made during the online re-planning.

The patient-specific component is especially important. Homogeneous or simplified anthropomorphic phantoms remain useful for routine machine QA, but they do not reproduce the geometric complexity of real anatomy or the tissue interfaces that are most relevant in high-field MR-guided radiotherapy. By generating phantoms directly from patient CT or MR images, the proposed method provides a way to build verification objects that reflect the clinical configuration of interest. This has potential value for site-specific QA, for end-to-end testing of adaptive workflows, and for development studies in which anatomically realistic test objects are needed before translation into clinical practice.

A second important feature is the integration of embedded dosimetry into the phantom-design process in which the holder design and insertion-path are incorporated during phantom generation. Minimizing traversal through anatomically heterogeneous regions is a practical design consideration, since it reduces abrupt material transitions and helps preserve the local imaging and dosimetric properties of the phantom around the measurement site. The ability to support OSLD, film, and ion-chamber configurations further increases the flexibility of the platform and suggests that the design can be adapted to a range of QA tasks requiring either planar or point measurements.

The material-characterization results also support the feasibility of multi-modal phantom fabrication. We find that calibration curves specifying CT or MR properties can be tuned through different components of the printing formulation, with radiopaque material content primarily affecting attenuation and gel/tissue mixtures influencing MR signal intensity. This allows flexible assignment of imaging properties to different anatomical structures rather than relying on a single uniform material response. Printed material robustness across both CT and MR was not investigated, but could be achieved through the creation of calibration curves for MR and CT produced from the simultaneous variation of GelMatrix, and RadioMatrix™.

The 4D extension of the workflow is another notable aspect of the study. Adaptive radiotherapy on MR-Linac systems is often influenced by organ motion, especially for thoracic and abdominal targets. The ability to generate phantoms across motion states and interpolate between them provides a pathway toward motion-aware QA objects that extend beyond static validation. In the current implementation, the method preserves mesh correspondence across phases and generates temporally continuous phantom representations from discrete input motion states. This provides an initial step in future work involving dynamic dose verification and accumulation for motion management strategies.

The presented workflow has several limitations. First, the quality of the final phantom depends on the accuracy of the automated segmentation and the quality of the input imaging data. Segmentation errors, missing structures, or image artifacts may propagate into the mesh-generation and fabrication steps. We recommend users review the mesh after this step in the process. Manual edits may be needed. Second, the dosimeter-placement step is intentionally simple and serves as a proof of concept. However, a more comprehensive and efficient search strategy could improve placement in anatomically complex cases. Third, while sample measurements generated calibration curves for a family of material mixtures, the long-term mechanical stability, repeatability, and radiation-response characteristics, such as MR relaxation of the printed materials were not fully evaluated in this study. Finally, the 4D framework currently addresses geometric motion representation; translation into a physically actuated moving phantom is work in progress.

Despite these limitations, the proposed workflow provides a practical starting point for constructing patient-specific phantoms tailored to MR-Linac QA applications. The design is modular and can be expanded as segmentation methods, printable materials, and fabrication methods continue to improve. Future work should include end-to-end dosimetric validation of adaptive treatment workflows, quantitative assessment of detector placement accuracy, evaluation of long-term material stability, and development of physically actuated implementations of the 4D phantom concept. This framework may help bridge the gap between digital QA methods and physical validation phantoms.

## CONCLUSIONS

We developed an end-to-end method for automated design of patient-specific, multi-material 4D-printed phantoms for quality assurance of adaptive radiotherapy on a 1.5T MR-Linac. The framework integrates patient-image segmentation, high-resolution mesh generation, dosimeter-holder placement, multi-material assignment, and motion modeling into a unified workflow. The results demonstrate feasibility of generating anatomically realistic phantoms with embedded dosimetry and tunable CT and MR imaging properties. In addition, the 4D implementation enables temporally continuous phantom representations with consistent topology across motion states, providing a foundation for motion-aware QA applications. This approach offers a practical pathway toward reproducible, imaging-compatible, and anatomically relevant physical phantoms for end-to-end validation of adaptive radiotherapy workflows.

## Supporting information

4D Phantom

## Data Availability

All data produced in the present study are available upon reasonable request to the authors.

## ACKNOWLEDGMENTS

The authors are grateful to Drs. Samuel Hellman and Justin Kuligowski for guidance with 3D-printing methods.

## APPENDIX A

**Table A1.** Relation between index (Ind) and material composition for the calibration curves presented in Fig. 6 in the manuscript. Number in front of each material represents percentage of corresponding material present in mixture. GM = GelMatrix, TM = TissueMatrix, BM = BoneMatrix, RM = RadioMatrix, AG = Agilus30, VW = Vero PureWhite

| Ind | Mixture | Ind | Mixture |
| --- | --- | --- | --- |
| 1 | 10RM-40AG-50GM | 1 | 10GM-90AG |
| 2 | 10RM-50GM-40VW | 2 | 20GM-30AG-50RM |
| 3 | 10RM-90AG | 3 | 30GM-70AG |
| 4 | 10RM-90VW | 4 | 33GM-33AG-33TM |
| 5 | 20RM-30AG-50GM | 5 | 50GM-50AG |
| 6 | 20RM-50GM-30VW | 6 | 20GM-80TM |
| 7 | 30RM-20AG-50GM | 7 | 60GM-40AG |
| 8 | 30RM-50GM-20VW | 8 | 40GM-60TM |
| 9 | 30RM-70AG | 9 | 60GM-40TM |
| 10 | 30RM-70VW |  |  |
| 11 | 40RM-10AG-50GM |  |  |
| 12 | 40RM-50GM-10VW |  |  |
| 13 | 50RM-40AG-10GM |  |  |
| 14 | 50RM-10GM-40VW |  |  |
| 15 | 50RM-20GM-30AG |  |  |
| 16 | 50RM-20GM-30VW |  |  |
| 17 | 50RM-30GM-20AG |  |  |
| 18 | 50RM-30GM-20VW |  |  |
| 19 | 50RM-40GM-10AG |  |  |
| 20 | 50RM-40GM-10VW |  |  |
| 21 | 50RM-50AG |  |  |
| 22 | 50RM-50VW |  |  |
| 23 | 70RM-30AG |  |  |
| 24 | 70RM-30VW |  |  |
| 25 | 90RM-10AG |  |  |
| 26 | 90RM-10VW |  |  |
| 27 | 100% RADIOMATRIX |  |  |

## APPENDIX B

**Table B1.**
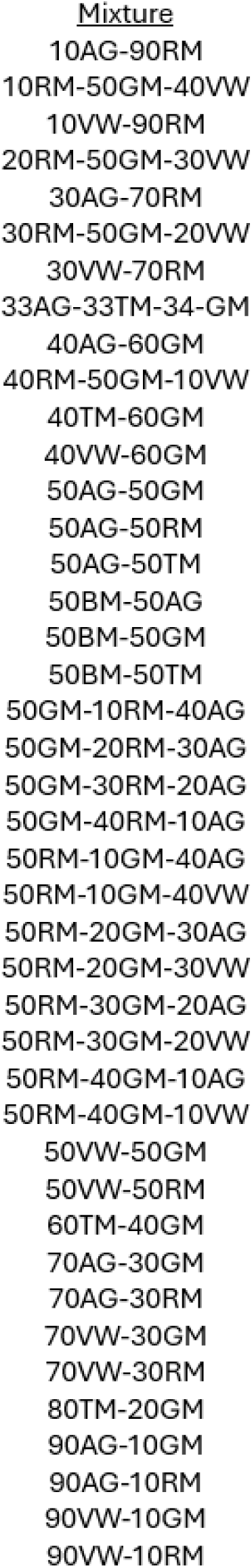
List of all material compositions included in the measurements for the calibration curves presented in Fig. 6 in the manuscript. Number in front of each material represents percentage of corresponding material present in mixture. GM = GelMatrix, TM = TissueMatrix, BM = BoneMatrix, RM = RadioMatrix, AG = Agilus30, VW = Vero PureWhite

| <u>Mixture</u> |
| --- |
| 10AG-90RM |
| 10RM-50GM-40VW |
| 10VW-90RM |
| 20RM-50GM-30VW |
| 30AG-70RM |
| 30RM-50GM-20VW |
| 30VW-70RM |
| 33AG-33TM-34-GM |
| 40AG-60GM |
| 40RM-50GM-10VW |
| 40TM-60GM |
| 40VW-60GM |
| 50AG-50GM |
| 50AG-50RM |
| 50AG-50TM |
| 50BM-50AG |
| 50BM-50GM |
| 50BM-50TM |
| 50GM-10RM-40AG |
| 50GM-20RM-30AG |
| 50GM-30RM-20AG |
| 50GM-40RM-10AG |
| 50RM-10GM-40AG |
| 50RM-10GM-40VW |
| 50RM-20GM-30AG |
| 50RM-20GM-30VW |
| 50RM-30GM-20AG |
| 50RM-30GM-20VW |
| 50RM-40GM-10AG |
| 50RM-40GM-10VW |
| 50VW-50GM |
| 50VW-50RM |
| 60TM-40GM |
| 70AG-30GM |
| 70AG-30RM |
| 70VW-30GM |
| 70VW-30RM |
| 80TM-20GM |
| 90AG-10GM |
| 90AG-10RM |
| 90VW-10GM |
| 90VW-10RM |

